# Natural is divine: Religious leaders’ nuanced views on birth spacing and contraceptives in Sierra Leone - qualitative insights

**DOI:** 10.1101/2023.10.06.23296669

**Authors:** Regina Yillah, Florence Bull, Alhaji Sawneh, Beryl Reindorf, Hamid Turay, Haja Wurie, Mary Hodges

**Affiliations:** Institute for Development (IfD), Freetown, Western Area, Sierra Leone; Christian Health Association of Sierra Leone, Freetown, Western Area, Sierra Leone; Njala University, Njala, Moyamba District, Sierra Leone

## Abstract

This research explored the viewpoints of 116 religious leaders in Sierra Leone including 32 Muslims and 84 Christians from nine different denominations. The study’s primary objectives were to understand their perspectives on family planning, modern contraceptives, sexual reproductive health education, and the religious doctrines influencing these beliefs. The study also aimed to gauge their knowledge of family planning and modern contraceptive methods.

In September 2021, data was collected from religious figures purposefully selected from 11 districts and the Western Area Urban through 16 focus group discussions. The discussions, initially conducted in local languages, were translated and transcribed into English. The data was then subjected to a thematic analysis using NVIVO 12 software.

The analysis revealed diverse opinions, both across different religions and within specific denominations. A common thread was the general support from both Christian and Muslim leaders for natural birth spacing methods, with some reservations about artificial techniques. There were clear distinctions in beliefs among denominations: Catholics largely considered artificial contraceptives to be against the divine will, while Pentecostals and some Muslims found them permissible under certain conditions.

The findings underscore the potential of religious leaders in Sierra Leone to act as influential advocates for family planning, given their support for natural birth spacing. To maximize the impact of advocacy efforts, the study suggests a focus on engaging Pentecostals and Muslim leaders rather than Catholics. Additionally, religious leaders with professional backgrounds in health or education appear more receptive to modern contraceptive methods and could be particularly valuable allies in these endeavours.

## Introduction

Globally, it is estimated that family planning (FP) can prevent up to one-third of maternal deaths [1], while birth spacing of at least 36 months can reduce under-five mortality by 25% [2,3]. High fertility is an important determinant of both maternal and child mortality and morbidity [4]. From 1990 to 2019, the global fertility rate declined from 3.2 to 2.5 children per woman and from 6.3 to 4.6 in sub-Saharan Africa [5]. In Sierra Leone, the total fertility rate declined from 6.6 in 1990 to 4.2 in 2019 [6.7] and the modern contraceptive prevalence rate (mCPR) for all women increased from 17.4 in 2012 to 26% in 2022 [8].

Sierra Leone has one of the highest maternal mortality ratios in the world (717 per 100,000 live births) coupled with high neonatal (31 per 1,000), infant (75 per 1,000), and under-five (108 per 1000) mortality rates, in 2021 [7]. This represents a significant improvement from previous years and is a testament to the efforts being made to improve health outcomes for this target group. However, there is plenty of room for improvement in addressing the underlying causes of maternal mortality, such as poverty, poor nutrition, limited access to FP services and women’s ability to make decision regarding their own health.

Family Planning (FP) as defined by the World Health Organization ‘allows individuals and couples to anticipate and attain their desired number of children and the spacing and timing of their births. It is achieved through the use of contraceptive methods and the treatment of involuntary infertility’ [9]. It is one of the single most important determinants of child survival, yet a quarter (25%) of women in Sierra Leone have an unmet need for family planning (32% for birth spacing and 14% for limiting births) [7]. The teenage pregnancy rate in the country is 34%, and teenage childbearing contributes to 40% of maternal deaths [8]. About 48% of young women (20-24 years of age) have become mothers before the age of 18 [10]. Early childbearing prevents girls from achieving their full potential, contributing to high school dropout rates and the consequently low status of women and reduced income generation [10].

Recognizing the importance of informed choices in reproductive health, the Sierra Leone Ministry of Basic and Senior Secondary Education (MBSSE) recently introduced a policy and curriculum for comprehensive sex education (CSE) and facilitation of access to reproductive health service delivery in schools. This CSE policy was developed to help young people make informed sexual and reproductive health choices, which is central in reducing teenage pregnancy, early child marriage and improving government and civil society responsiveness to women’s and girls’ needs [11].

Universal access to FP is central to gender equality and women’s empowerment, and a critical factor in reducing poverty. It reinforces women’s right to determine the spacing of their children and ultimate family size. In developing countries, millions of women want to delay or avoid pregnancy but do not have access to or are not using an effective method of FP. In sub-Saharan Africa, 58 million women have an unmet need for family planning, and within the region, the 39 poorest countries’ unmet need for family planning has increased since 2008 [11]. Addressing the unmet need for family planning requires a multi-faceted approach that increases access to education, information, and healthcare services for women and families, and includes addressing cultural and social norms as well as religious beliefs that often deter uptake of modern contraceptives.

Most women in Sierra Leone know that good quality, modern contraceptives are available in their communities, yet choose not to access it or to discontinue its use [7]. Religious belief is a key factor affecting women’s uptake and continuation of modern contraceptives; 9.3% of women discontinue family planning based on religious grounds [7]. Studies have shown that religious leaders command respect in their communities, and community members are likely to listen to them about family planning and the use of modern contraceptives [13]. They are strategically placed to either promote or deter uptake of modern contraceptives.

A systematic review seeking to clarify the role of religion and religiosity on fertility and contraceptive use in continental Sub-Saharan African countries found the following: 1) Islam followers have higher fertility rates than followers of Christianity (Catholics having the highest fertility among Christians, followed by Protestants and Apostolic) 2) Religion exerts an influence on fertility rates and contraceptive use, but the magnitude and nature of this influence can differ significantly from one country to another 3) Within the context of Islam a stronger degree of religiosity and higher fertility rates correlate with decreased contraceptive use, unlike among Christians where this is not the case [14].

This study, therefore, sought to explore the religious leaders’ perspectives on the family planning methods and contraceptive use among women of childbearing age including their knowledge of the different methods, what they believe their religion says about family planning and contraceptive use; and the Quranic and Biblical references they use to justify their views. It also sought to examine their views on reproductive and sexual health education and to identify champions and entry points for family planning. This knowledge will provide policymakers with critical insights into potential entry points for engaging with religious leaders. Understanding how to leverage the influential positions of these leaders can enhance the effectiveness of family planning advocacy and influence the decisions made by individuals and couples.

## Materials and Methods

This qualitative study used focus group discussion (FGD) to allow flexibility for respondents to explain what they knew to be important, the intricacies of the issues, their moral values and religious beliefs.

Purposeful sampling allowed the researcher to recruit participants that were knowledgeable about either Islam or Christianity. More Christian religious leaders were recruited as the study was commissioned by the Christian Health Association of Sierra Leone (CHASL) whose primary interest was understanding the perspectives of Christian religious leaders, while a comparison of their views with Muslim leaders was of secondary interest. One FGD was conducted with 6-8 religious leadersin each district headquarter-town. To avoid contention, we held FGDs with Muslim and Christian scholars separately.

An FGD guide that included the study information, consent procedures and a participants’ demography sheet was used. The guide included open-ended questions and probes on knowledge and perception of family planning, modern contraception and references from the Quran and Bible, as shown in Figure 1. The guide was developed in English and translations of key phrases in all the main local languages were agreed on with data collectors and documented during training. The FGDs were conducted in either Krio, Temne, or Mende, depending on the preferred local language of the respondents. In-depth responses were obtained about participants’ thoughts and feelings to gain insights into their beliefs and attitudes towards FP services. Data collectors took notes in English and the interviews were recorded and translated and transcribed to English by two native speakers of the local language.

Recordings were stored in an encrypted telephone device and later transferred to the secure OneDrive. All audio transcriptions were also stored in the same drive and only shared with the study team.

All FGDs were audio recorded to ensure accuracy and completeness of the collected information, and supplementary notes were taken to capture immediate insights and contextual details. The audio recordings from the FGDs were listened to and directly translated into English transcripts. These transcriptions were then cross-referenced with the written notes to ensure accuracy and completeness of the data. Responses were analyzed using NVivo 12 software to synthesize results into major themes using thematic content analysis.

## Results

A total of 116 religious leaders participated (Muslims: 32, Christians: 84) in September 2021. 84 Christian religious leaders, representing nine different denominations, were included in the study. The breakdown of Christian denominations are shown in Table 1. Most of Christian participants identified as Pentecostal, accounting for 38.1% (n=32) of the participants. Methodist (14.3%, n=12) and Wesleyan (13.1%, n=11) were the next most common denominations, followed by Baptist (10.7%, n=9). The Anglican denominations represented 6% of the sample (n=5) Catholic and Evangelical denominations each represented 7.1% (n=6, respectively) of the total participants. Protestant participants were relatively few, accounting for only 2.4% (n=2) of the total. A single participant identified as non-denominational, constituting 1.2% of the total participant population.

**Table 1:** Christian Denominations.

| Denomination | Number of Participants | Percentage of Total |
| --- | --- | --- |
| Anglican | 5 | 6.0 |
| Baptist | 9 | 10.7 |
| Catholic | 6 | 7.1 |
| Methodist | 12 | 14.3 |
| Evangelical | 6 | 7.1 |
| Non-denominational | 1 | 1.2 |
| Pentecostal | 32 | 38.1 |
| Protestant | 2 | 2.4 |
| Wesleyan | 11 | 13.1 |
| **Total** | **84** | **100** |

Thirty-two Muslim religious leaders participated, but their denominations or sects were not noted as the main purpose of their participation was to contrast their views with those of Christian religious leaders. It should be acknowledged for contextual relevance that the majority of Muslims in Sierra Leone predominantly identify with either the Shia or Sunni branches of Islam.

The results have been categorized into religious leaders’ knowledge of contraceptive use and birth spacing; perceptions of modern contraceptives; perception of sex education; and identifying champions and entry points for FP advocacy and service delivery.

### Religious leaders’ knowledge of family planning methods

The results show that religious leaders have a fair understanding of FP and the different types of modern contraceptives. They categorized FP methods into 1) Artificial/modern contraceptives that include pills, injections (Depo-Provera), and implants (locally known as Captain Band), intrauterine devices (coil), male and female condoms and 2) natural and traditional methods such as withdrawal, calendar method (i.e., menstrual cycle tracking to avoid sex during the fertile days), abstinence, and lactational amenorrhea method. They stated that the natural methods are safe and advantageous as they have no side effects.

The analysis revealed varying opinions between Christian denominations and Muslims about family planning and use of modern contraceptives. However, an area of concurrence was that all Christian and Muslim religious leaders fully support the practice of birth spacing and could outline associated benefits. For example, ensuring the woman has good reproductive health and time to take care of the children with the limited resources available, instead of having “*wan na belle, wan na back” (one in the belly and one on the back – a local slogan for inadequate spacing) Wesleyan* Christian, Western Area Urban.

In the exploration of religious leaders’ perspectives on family planning, several themes emerged that demonstrated an in-depth understanding of the diverse benefits associated with family planning. The succinct but profound statements encapsulate the various dimensions of family planning, encompassing not only its health benefits but also its wider socioeconomic implications.

*“It saves lives and improves the health of women that have given birth. It also allows parents to take care of their homes and time for children to grow properly before another pregnancy. Family planning also prevents girls from dropping out of school and achieving their goal”* Catholic Christian, Western Area Rural.

*“The relevance of spacing birth is for instance, if you have space for your children for more than three years, the older children will be able to take care of the younger ones, and the parents too will be able to cater for their children. These are the things we preach to our congregation about the importance of birth spacing.” –* Muslim, Falaba.

Another Muslim leader cited a recommended breastfeeding duration of 30 months, which surpasses the World Health Organization’s recommendation of breastfeeding “up to the age of two years and beyond.” This extended breastfeeding period aligns closely with the three-year birth spacing mentioned by other participants in the study.

“*To me, giving birth is very important because it’s planned by God. Allah said we should marry and give birth to children, and the Quran said (2:223) when a woman is pregnant, she should breastfeed for at least 30 months, if we follow this, it will be okay*.”- Muslim, Kailahun Overall, religious leaders voiced that birth spacing is acceptable, however as shown below they less open to the concept limiting births.

### Religious leaders’ views on use of contraceptives for limiting births

Limiting births refers to individuals or couples who have completed their families and do not plan to have any more pregnancies. Religious leaders shared that children are from God and any means of limiting their existence is prohibited and not of God’s ways, as stated:

“*Psalm 127- the child is a heritage from God, and we believe in procreation, so anything that destroys it, like the contraceptive, is not of God and it is against the Christian beliefs*, *and Genesis 1:28 states “be fruitful and replenish the earth.”* – Catholic Christian, Port Loko

Others Christian leaders especially those from Evangelical and Protestant denominations held contrary views; they stated that even though the Bible (Genesis 1:28) states children are a blessing from God and that people should be fruitful and multiply, they did recognise that these ’commands’ should not be taken literally as the word of God is allegorical.

“*The commandment of God in the Old Testament is we should be fruitful and multiply. By looking at the perspective of fruitfulness and multiply, at that time, there were not enough people on planet earth, now, there are lots of people on planet earth, and some people are complaining that we are overpopulated. Instead of having so many children, one or two will be enough. By looking at the current socio-economy, a bag of rice costs New Leones 370-500 (∼$40-$50). If you give birth to 30 children, how many days will it take to finish a bag of rice*?” – Pentecostal Christian, Koinadugu.

Another Pentecostal Christian Leader suggested that not having the means to take care of your children is also a sin and, thus, the importance of having financial security before deciding to have children.

“*According to Pentecostal, which is my denomination, it preaches against bearing children and leaving them unattended… that is a sin. Also, failing to provide the fulfilling demands of your children is not sinful but criminal. This is what our religion (Pentecostal) says about childbearing and family planning.*” – Pentecosal *Christian, Bonthe*

The Pentecostal Christian beliefs above differed significantly to those of Muslims who stated that Islam is against family planning if the intention is due to fear of provision for and income to take care of the children, then it is haram, as the Quran 17:31 states:

“*Kill not your children for fear of want: We shall provide sustenance for them as well as for you. Verily, the killing of them is a great sin*”. Muslim, Western Rural.

Overall, the Islamic perspective was that family is acceptable if done following Islamic practice (for example 30 months of breastfeeding) and with a good intention of safeguarding the woman’s health, however it should not be done out of fear of not being able to provide for the family.

Some Muslim respondents also stated that the use of modern contraceptives is a way of aborting a child, which is against the teaching of Islam. Abortion is a sin. However, this view was not universal among Muslim participants.

Finally, Catholic Christians expressed that while they were in favour of family planning both for spacing and limiting, their main concern was the means by which family planning is achieved – the use of modern contraceptives which are artificial and have many side effects.

### Perceptions of modern contraceptive methods

The Catholic Christians leaders stated that the Bible only supports natural FP, and anything that suppresses procreation artificially goes against the word of God.

“*At the Catholic Church …. we accept family planning, but we are against the methods that are being used. Of course, the natural way is natural, and there is no intervention from a human being. The others are artificial.*“ Christian, Western Area Urban

A Muslim leader also stated that during the time of the Holy Prophet, his disciples used the withdrawal method, and when the prophet was asked about it, he said, “Not out of all the semen a child is formed, and if Allah willed to create something, nothing would stop Him from doing it [15].” This indicates willingness on the part of Muslims to accept family planning using natural methods.

Christian religious leaders of all denominations and Muslim religious leaders also voiced concerns about side effects, infertility, and the professional capacity of health workers. One of the primary concerns expressed by several Christians was the negative effects of modern contraception; these adverse effects include difficulty conceiving, stomachache, infertility, severe internal bleeding, blockages, cancer, irregular menstrual cycle, and changes in blood pressure.

“*Some disadvantages are like when someone has taken the pills or injectables, they are faced with a lot of risks, and they would affect your blood pressure because there are hormones that have been injected into your system that may affect you negatively*.” – Methodist Christian, Western Urban.

Some of the religious leaders mentioned side effects of women having big stomach bloating and/or abdominal growth or fibroid and disruption of their menstruating cycle. They believe that such bloating affects their beauty and will lead to them being barren or infertile. Concerns about cancer from intrauterine devices (IUDs) and the possibility of the IUD entering into the stomach, causing excess bleeding to death.

*“Sometimes this IUD they use may cause abortion, and this may happen when the coil gets displaced into the stomach while the woman gets pregnant at the same time. This would lead to abortion because there are two conflicting foreign bodies competing for space, so one has to be displaced, leading to miscarriage.” - Catholic* Christian, Western Urban.

Christian leaders’ additional concerns about FP include condom getting stuck and condom tearing during intercourse, putting one at risk of sexually transmitted diseases and unwanted pregnancy. Another serious concern is out-of-pocket expenses resulting from side effects and complications. Furthermore, they assert that not all health workers have the capability to provide high quality counselling and modern contraceptive administration.

*“Some nurses cannot insert the implant properly, causing bleeding and unrest”,* Catholic Christian, Western Urban.

Another major concern raised was the perceived indiscriminate dissemination of modern contraception to school-aged children in schools. They believe that access to contraceptives encourages sexual activity, urging children to test out what they have learnt about. They suggested this practice will eventually destroy the lives of children, causing them to be disobedient to their parents. They assert that providing contraceptives to underage children without parental consent is inappropriate.

“*Government give children, even the underage, without any parental consent. This has negative implications on the children …as parents, we see some it has some health effects on them. My child is an example of it; they gave her an implant without my notice. I was made to understand about it the day she got an attack which left her unconscious.”* – Baptist Christian, Port Loko.

The influence of modern contraceptives on children was another critical concern. Some believed that modern contraceptives are making them promiscuous and causing them (particularly girls) to get loose in the streets. Furthermore, modern contraceptives divert students’ attention away from their studies and allow them to disregard preventative measures and the fear of God or become wayward.

“*Family planning is like giving our wives and daughters licenses for promiscuous behaviour; as a result, the women and/or daughters become unruly and disrespectful to their husband*“. Muslim, Falaba.

However, some argued that FP is good and, if done correctly, can lead to positive outcomes such as ensuring a prosperous future for children by allowing them to gain an education and support. Additionally, they stated that FP is good for school-going children and society as it assists in the prevention of teenage pregnancies and allows girls to reach their potential.

### Identifying champions and entry points for family planning

The findings indicate that there are religious leaders (both Christian and Muslim) who advocate for the use of modern contraceptives because of their professional lives, working as nurses or teachers. They are key influencers who can raise awareness of FP, therefore making them potential champions for FP in their communities.

Religious leaders mentioned that they are aware of their potential role to improve sexual and reproductive health. However, some stated that they had not been involved enough to take up the dissemination of messages at the community level.

*“We are always with the people, and they listen to us. Therefore, Government should involve us more in more complicated issues like FP to be the community ambassadors, [and] then you will see how the uptake will improve”* Muslim, Western Rural.

Religious leaders further mentioned that they have several ways of reaching many people to promote FP practice.

### Preaching family planning to the congregation

Muslim leaders stated that they sometimes preach about FP while Catholics believe that FP is not a suitable topic to preach in the church and that it can cause division in the church. On the other hand, Pentecostal Christians were willing to preach about FP, especially the natural methods, as long as it does not promote promiscuous behaviour. The Pentecostal added that preaching about FP is not always done from the pulpit because the congregation is a combination of children and adults.

“*…for instance, in a congregation, sometimes we as pastors do find it difficult to go too deep to talk about the uses of these contraceptives that we have been discussing because we have young people in our midst, and as a pastor, we know that family planning is good, but we have to discuss that with people that are married and also the uses of these contraceptives. We just cannot explain that in the church where we have young people that are not married, that will want to prompt them to fornicate, which is not good.*“- *Christian*, *Kono*

The Muslim respondents added:

*“There is a subject that we call ’fedarr’; it is one of the materials we use to preach about family planning, its usefulness and how to go about it. We have another subject which is called Lull; Lull tells them how to use family planning and also how to go about it. We treat those two subjects.” – Muslim, Pujehun*

The respondent also mentioned that they preach to unmarried youths that are sexually active to use contraceptives for them to continue schooling and prevent them from giving birth out of wedlock, especially for ruling/chieftaincy households. We told them that FP is good using the famous quote, “if you plan yourself, it will be beneficial. If you fail to plan yourself, it’s not good.”

*“They should join family planning as it will prevent you from giving birth to a child out of wedlock because, in the ruling house, children out of wedlock will not be eligible to participate in any chieftaincy activities. It will be a disgrace to the child in future; therefore, we encourage them to use family planning to prevent such awkward scene happening” - Muslim, Pujehun*

### Discussion about family planning in different religious groups

Some churches, for example, the United Methodist Church, have a policy where they allocate some time for health talks before each sermon on various topics.

“*After Ebola Virus Disease, the United Methodist Church has made it a policy for anyone who wants to mount the pulpit to give a health talk for 10 minutes before preaching the actual sermon. Before you go to the pulpit, we sit together and agree on a particular health talk; that is the time we address our members about family planning. Little by little, the church is getting acquainted with lots of health information.*“-*Christian, Moyamba*

The Pentecostals also have a platform called RETREAT which they utilise to preach to their members about FP.

“*Yes, at Pentecostal, we preach to our members about family planning during service or through a platform called RETREAT. This a platform where we assemble our youths-young adults, women’s wing. There are different sectors of retreats. The singles retreat is meant for those that are not married, where they are taught about restraining or bearing a child for someone who has not tied the knot with you. There is another retreat called couples’ retreat, whereby married couples come together and listen to advice delivered. “*- *Christian, Bonthe*

Religious assemblies such as youth wing, women’s and men’s groups, and persons with disabilities also discuss FP. The main topic discussed is abstinence for unmarried youth. They should maintain celibacy as sex is for married people, which implies that FP is for married couples. In addition, the youths are advised about moral values and religious positions on FP and/or health practitioners are invited to discuss this and other different health topics.

### Religious leaders’ perspective on reproductive and sex education

The majority of Christians (all denominations) supported sex education in schools and believed in the new school curriculum that integrates comprehensive sex education. They understood the adverse effects of not being well informed about reproductive health and sex, including unwanted pregnancies, which could be detrimental, especially to the life of the girl child and their parents.

“*Yes, my church supports that because it is a burden to the child and the parents when a child bears a child when she has not attained the age and the capacity. This may lead to dropout in schools because here in the provinces, when a child gets pregnant, the parents consider that as the end of her education, and they would never care about her continuing to school again.*“– Christian, Bombali

The minority of Christian leaders who opposed sex education in schools expressed that if children become aware of sex and are supplied with modern contraception, it reduces children fear of becoming pregnant, making them more sexually active with multiple partners, losing focus in their education, boys taking drugs for longevity, becoming thieves and drug addicts. On the other hand, the Islamic position was that sex education is not suitable for children; rather, such information should be provided to adults instead of children.

*“It’s not good. Because that is the main reason, underage girls are becoming pregnant nowadays. Even as parents, we are doing it secretly. That’s not good; if they were mature, it’s different. But underage children will lead to doing bad things. If they are above 18 years, that’s not bad. This has brought so many problems because these children are now practising it, and it’s not good at all.” Muslim, Kailahun*

## Discussion

Religious leaders’ knowledge of modern contraceptive methods was similar to the rest of the population, where 98% of married women and 99% of men know at least one method of family planning [7]. Their understanding of FP is in line with religious leaders across the globe that describe FP as a means of voluntary prevention or spacing pregnancy [16].

Both Muslim and Christian religious leaders supported birth spacing – this is an important point of agreement between with FP proponents. It provides an entry point for influencing religious leaders to be more supportive of FP. However, there are distinct barriers to cross with different religious groups and denominations, and we set out our contemplations on these barriers below. Some Muslim leaders have stated that modern contraceptive is permissible only if there are health implications (i.e. if there is a high chance of losing either the mother or the child’s life during pregnancy or delivery) [17]. A study in Burkina Faso also revealed that religious leaders believe that modern contraceptive methods are against the principles of religion [18]. These contrasting viewpoints indicate a complex interplay between religious doctrine and the acceptability of modern contraceptive methods. While some Muslim leaders see a space for modern contraceptives in specific health-related circumstances, other religious figures, as evidenced in Burkina Faso, remain firmly against their use on religious grounds. This divergence suggests that religious teachings on contraception are not monolithic but may vary significantly both within and across religious communities, thus presenting both challenges and opportunities for family planning advocacy.

Catholics believe that the only way to achieve birth spacing is through natural methods. Natural FP methods are known to be less effective than modern contraception. However, religious leaders argued that modern methods come with many side effects, whereas natural methods do not. A study in Kenya, Nigeria, and Senegal, reported side effects and health concerns such as “long-term infertility”, “safety reason”, “end up with health problems (48–74%)”, “dangerous to women’s health (47–72%) and “contraceptives can harm your womb (37–62%)”) as the main reason for non-use of modern contraceptive [19].

This tension between the effectiveness of modern contraception and the perceived safety of natural methods presents a complex landscape for policymakers and family planning advocates.

While the Catholic stance may limit the uptake of highly effective modern contraceptives, it also opens a door for compromise by promoting natural family planning methods to a sector of the population that might otherwise avoid birth control altogether. Given the religious leaders’ concerns about the side effects of modern contraceptives, there is an opportunity for more nuanced conversations and educational programs. These could focus on debunking myths and providing evidence-based information about the safety profiles and side effect management of modern contraceptives. Moreover, this approach can help create a common ground to engage religious leaders as allies, allowing them to become more informed and possibly more receptive to other contraceptive methods over time. In essence, a tailored, evidence-based advocacy could serve as an entry point for engaging with religious communities that have reservations about modern contraceptive methods.

Pentecostal Christian leaders, on the other hand, appeared less doctrine-led and are more practical. They paid attention to economic factors highlighting that people should have the children they can afford. They then extrapolate that to say that having children that you cannot afford is a sin, but this is not backed by any biblical quote. The pragmatic orientation of Pentecostal Christian leaders toward family planning suggests that they may be more amenable to conversations about the implementation and promotion of modern contraceptive methods within their congregations. Their focus on the economic aspects of family planning, although not explicitly grounded in religious doctrine, provides a crucial avenue for engagement. The absence of doctrinal rigidity may serve as a more accessible entry point for policymakers and family planning advocates to present evidence-based information. The key to effectively engaging with Pentecostal leaders would be to frame the conversation around their existing concerns. Highlighting how modern contraceptive methods can be both safe and economically beneficial may help align their religious messaging with broader public health goals. Evidence-based discussions about the safety of modern contraceptives and options for side effect management could be particularly persuasive in this context. Such an approach would not only facilitate the religious leaders’ endorsement of family planning but could also lead to active advocacy, thereby amplifying the reach and impact of family planning initiatives.

Some Muslim leaders stated clearly that fear of responsibility is not a reason for limiting births. Rather they believe that it is only acceptable if it will affect the health of the women. For Muslim leaders, therefore, the key messaging has to focus on the health of the woman. They are not in principle against artificial methods – so they could be influenced to promote modern FP methods through a birth spacing lens. There were also some gender considerations; according to some Muslim leaders, contraceptive prevention is permissible only if the husband approves, and using different methods is not a problem as long as it is not with the intention of permanent effect. Any method that causes permanent sterilization without any proper medical reason is prohibited or haram. This illustrates clearly that it would be difficult to influence Muslim religious leader to promote permanent methods such as sterilization.

The position articulated by some Muslim leaders indicates complex yet promising entry points for public health practitioners and policymakers who aim to promote family planning. The leaders’ openness to birth spacing using modern contraceptive methods, if framed within the context of women’s health, provides a strategic avenue for engagement. Given that their religious beliefs do not wholly reject the use of artificial methods, Muslim leaders could serve as potential allies in normalizing the conversation around modern contraceptives in their communities. However, the conditions set forth by these Muslim leaders also present specific challenges. The requirement for spousal approval and the prohibition against methods leading to permanent sterilization underscore the sociocultural and religious constraints that need to be navigated carefully. These insights imply that advocacy and education should not only be aimed at the leaders themselves but must also engage the broader community, including men, to build a consensus around the health benefits of family planning.

Furthermore, considering that permanent sterilization methods like tubectomy or vasectomy are ’haram’ (forbidden) without a substantial medical rationale, any policy or program targeting Muslim communities in Sierra Leone would need to be explicit about the reversible nature of most modern contraceptives, or otherwise focus on those methods that are temporary and reversible.

A major challenge to overcome with Muslims, however, is the education of adolescents on sexual reproductive health. The education of adolescents on sexual reproductive health among Muslim communities presents a unique dilemma, requiring a delicate and culturally sensitive approach. It’s important to reframe the conversation around the objective of such education: empowerment through knowledge. Family Planning (FP) program developers will need to make a compelling argument that the education provided is not a permissive gateway to sexual activity but rather an essential element of life education. Educating adolescents on how their bodies work can instill a sense of responsibility, bolster self-esteem, and equip them with the tools to make informed choices as they mature. This aligns well with values that are universally upheld across religious affiliations, such as personal responsibility and the preservation of human dignity. Hence, finding common ground in shared values could be a promising entry point for engaging Muslim religious leaders on the importance of comprehensive sexual reproductive health education for adolescents.

For all religious leaders, their concerns and misconceptions about artificial methods interfering with the natural processes that God has provided must be addressed. Furthermore, their concerns that modern contraceptives have side effects and lead to health problems and that providers are not well trained to provide the services will need to be addressed. Religious leaders’ knowledge and acceptance of birth spacing due to health risks of poorly spaced pregnancies can be an entry point to provide guidance on modern contraceptive use and debunking misconceptions. This will be the basis on which they can be encouraged to promote FP, thereby helping to reduce preventable deaths of mother and child. They could develop sermons/ Khutbah on ’family health and well-being’ with references from the Bible or the Quran that can use to address FP issues amongst their congregations.

Potential champions within the religious community are those who, in addition to being pastors or imams, also have other career roles, such as teachers or nurses. These dual-career individuals, because of their education or health professional training, understand the need for reproductive health education and the important role it plays in the reduction of teenage pregnancies, maternal and child mortality. However, their technical knowledge could also position them well to make informed arguments against family planning.

## Study limitations

The Christian Health Association of Sierra Leone (CHASL) commissioned the study, assisted in the study design and recruitment of participants. As a result, more Christian religious leaders were recruited as the primary interest was understanding their perspectives compared to their Muslim counterparts was of secondary interest.

## Conclusion

The understanding of family planning among religious leaders in Sierra Leone is largely reflective of the wider population’s views, presenting both challenges and opportunities for policy intervention. Given the pre-existing acceptance of ’natural’ methods for birth spacing within this cohort, there lies a significant opportunity to cultivate these leaders as champions for family planning. They could serve as influential change agents in promoting both the initiation and sustained use of family planning services within their respective communities. While the data suggests that Pentecostal and Muslim leaders may be more amenable to modern contraceptive methods compared to their Catholic counterparts, a multi-pronged approach will be required to engage each religious group effectively. Leveraging their acceptance of ’natural’ methods as a starting point could provide an entryway for more comprehensive discussions around modern family planning options.

## Data Availability

IfD archives all FGD guides and transcripts on site, which will not be shared to maintain confidentiality of respondents.

## Acknowledgements

The authors would like to thank all the Religious Leaders including FOCUS 1000 ISLAG and CHRISTAG for their collaboration during the field survey. Our appreciation to all the field workers for their cooperation and patience during the fieldwork

## Availability of data and materials

IfD archives all original FGD guides and transcripts on site, which will not be shared to maintain confidentiality of respondents.

## Ethics approval and consent to participate

Ethical clearance was obtained from the Sierra Leone Ethics and Scientific Review Committee at the Ministry of Health and Sanitation. The optional nature of the study (that participants could refuse to answer questions if they were uncomfortable, that from the study at any time in which case none of their data would be used) was explained verbally. All the respondents granted informed written consent to participate, and all data was anonymized. Confidentiality was also assured.

